# Long-term within-person variation of routinely measured biomarkers are associated with mortality and cardiovascular health

**DOI:** 10.64898/2026.05.04.26352236

**Authors:** Anthony J. Webster, Cynthia Wright Drakesmith, Rafael Perera-Salazar, David Steinsaltz, COMPUTE team

## Abstract

Biomarker measurements can assist with disease diagnosis and the assessment of disease risks, with the most recent measurements usually used by disease-risk models. However, a growing number of studies suggest that in addition to a biomarker’s value, its inherent variability, estimated from several measurements over many days or years in an individual, can convey independent prognostic information about disease risks. Variance estimates require an individual’s biomarker data to have been measured a sufficient number of times, ideally across a long time period, and are usually only available in a hospital setting or clinical trial. Furthermore, a single biomarker measurement will involve a combination of measurement-error, natural short-term variation over a daily time-period, variation over time periods of weeks and months, and slower age-dependent changes over several years. This paper develops a statistical method that accounts for these latter concerns, and applies it to Clinical Practice Research Datalink (CPRD) data collected by UK General Practitioners. It studies the associations between cardiovascular health outcomes and the within-person variances of eight routinely measured biomarkers. This involved Sequential Monte Carlo modeling to convert an individual’s biomarker measurements (collected over months or years), into estimates for the biomarker’s mean, linear age-dependent slope, within-person variance, and a variance due to variation on a daily time period or measurement errors. The result is a proof-of-principle that UK primary care Electronic Health Records (from CPRD) can be effectively used for this purpose. After adjusting for mean biomarker values, clear associations were found between mortality or cardiovascular disease risks and within-person variances for 6 of 8 biomarkers.

## Introduction

A large part of epidemiology involves the estimation of associations between disease risks and biomarkers such as body mass index (BMI) or blood pressure. Typical studies recruit a cohort of people, measure their biological characteristics, and then follow-up with records of disease diagnoses and the age at which they were diagnosed. As a result, most epidemiological studies use the value of biomarkers from a single time-point (at the study start). However, electronically linked health data are making it increasingly easy to obtain biomarker measurements from several time points in a person’s lifetime. This offers the potential for improved statistical assessments, but requires new methods to make best use of the incomplete and irregularly sampled data.

Biomarker variability has been most widely studied in the context of blood pressure [1, 2]. A 2023 review and consensus statement on the use of blood pressure variability in predictive studies [3], identified ten issues that a blood pressure model needs to mitigate or account for (labelled *(1)* to (10) in the text below):

I. For associations between blood pressure variance and disease risk, either scale the variance by the mean (a coefficient of variation), or include the mean as a covariate *(1)*.
II. Account for any potential long-term trend *(6)*.
III. Account for variance due to noise *(7)*, day-night variability and trends *(3-6)*, and the time between measurements *(5)*.
IV. Recognise that there are different blood pressure variances, that are defined by the timescales considered, and there is evidence that these have different associations with disease risks *(8)*. Here these points will be accounted for through: scaling the biomarkers (“standardising”), a “slope” describing longer-term trends, a within-person variance that models biological trends that occur over a timescale of weeks, and a “noise” variance to account for transient fluctuations on timescales of days or due to measurement error. The time between measurements are accounted for by allowing the typical size of changes in biomarker estimates to increase with time between measurements. Additional suggestions included [3]:
V. Including diastolic blood pressure variance, in addition to systolic blood pressure variance (multivariate modeling) *(2)*. For the proof-of-principle study considered here, we aim to determine the potential importance of biomarker variability for predicting disease risk, and to establish whether it can be estimated with routinely measured biomarkers. Therefore here we consider biomarkers independently of each other (with multiple adjustment for established risk factors). A further concern of [3] was,
VI. Due to the changing age and exposures of a patient, measurements for an individual’s blood pressure variation may not be reproducible *(9)*, leading to uncertainty over the prognostic value of the measurements *(10)*.

The biomarker model used here should improve the reproducibility of estimates by allowing for a long-term trend (slope), and accounting for the random variation over short time-scales as being an additional variance that is distinct from biological within-person variance. Despite these concerns, since the 2023 review numerous associations have been found between blood pressure variation and diseases. These include cardiovascular disease [4–6], stroke [7], renal disease [8], retinopathy [9], frailty [10], depression and anxiety [11–13], cognition [14, 15], and white brain matter [16– 18]. The quantity of associations and the existence of hypothesised causal mechanisms [19–22], provide a growing confidence that blood pressure variation is a biomarker for disease risks. A 2025 review [23] provides a comprehensive overview of present understanding of blood pressure variability and its associations with a range of diseases, and there is a 2025 meta-analysis that includes many of these studies [24].

There have also been several related studies that associate disease risk with the variability of other biomarkers that are considered here, these include: *low density cholesterol (LDL)* and cardiovascular disease or mortality [25–28], *pulse* variability and cardiovasular disease [27, 29], atrial fibrilation [22], stroke [30], liver disease [31], cancer [32], frailty [33], mental disorders [34], *sodium* variability and patient outcomes [35–39], *potassium* variability and patient outcomes ^1^ [40–45], *CRP* variability and cardiovascular disease [46], cognition [47, 48], diabetes [49], *UrACR* variability and changes in kidney function [50]. A limitation for many of these studies is the use of data from a hospital setting, which prevents the results from being applied by General Practitioners (GPs) in primary care, where the patient’s health and the available data are very different.

This article advances previous work in several ways:

1. We use routinely collected data in primary care electronic health records (EHRs) from the United Kingdom (CPRD Aurum), as opposed to data collected while in a hospital.
2. We demonstrate that the same method can be applied to a range of biomarkers - systolic blood pressure (SBP), diastolic blood pressure (DBP), resting pulse, total cholesterol to high density cholesterol ratio (TC/HDL), potassium, sodium, C-reactive protein (CRP), and urine albumin to creatinine ratio (UrACR).
3. Each trajectory is characterised by a mean, a mean gradient (or “slope”) that is intended to capture longer-term changes, and two variances. The variances are composed of a “noise” that is a combination of measurement error and short-term daily biomarker variation, and a within-person variability that describes transient intermediate-term biomarker variations.

This approach is intended to strengthen any associations with genuine biological variability by reducing regression dilution bias caused by errors in the estimated parameters, and to reduce the influence of long-term trends on estimates for standard deviations (that would otherwise be overestimated due to the changing biomarker levels). The model also aims to improve variance estimates by allowing and accounting for irregular individual measurement schedules, with greater changes expected with a larger time period between two measurements.

As explained earlier, the model is intended to account for many of the methodological challenges with using blood pressure variability as a covariate for assessing cardiovascular health, that were identified in a 2023 review and consensus paper on the importance of blood pressure variability

[3]. Because we estimate the long-term variation in biomarkers that are routinely measured by UK primary care doctors (for the purpose of assessing risk or assisting with a diagnosis), the approach has the potential to be implemented within routine clinical practice (see Discussion).

## Method

### Dataset and hospital diagnoses of disease

The cohort included over 6 million people, with data obtained from the Clinical Practice Research Datalink (CPRD) Aurum database [51], that contains anonymised health records of patients from a network of General Practitioner (GP) practices across England. The data were collected at GP practices, from adults that were continually registered for a minimum of 2 years between 2005 and 2021, and linked to hospital episode statistics (HES) and mortality data from the office for national statistics (ONS). Data includes regularly recorded biomarkers such as smoking status and body mass index (BMI), and test results that a doctor may have ordered for the purpose of diagnosis or monitoring a patient. The dataset was cleaned by the Primary Care data team to exclude a small proportion of data (less than 2 percent), that were for example, lacking a date or outside of a plausible biological range. Individuals from the UK regions of “South East” or “Yorkshire and the Humber” were removed to provide a test cohort, leaving approximately 5 million individuals. The hospital episode statistics (HES) include dates of episodes and disease diagnoses, that are coded with the International Classification of Diseases codes version 10 (ICD-10) [52]. For hospital-diagnosed diseases we consider the primary reason for admission (the primary diagnosis), that is the main reason for admission. This ensures that the disease has passed a threshold of severity to warrant treatment. Secondary diagnoses that are made during a stay in hospital may be of less severe disease, and are not considered here when identifying disease onset.

### Cohort exclusions and censoring

Individuals were excluded if they were aged less than 45 years or more than 84 years at the study’s start. Two age groups were considered, where the majority of potentially preventable disease occurs: middle age (45 to 64 years), and transition to old age (65 to 84 years). We included ethnic groups of Black, White UK, and Asian (excluding Chinese, for whom disease risks were found to be different, but were too few in number to allow their study as a separate ethnic group). The remaining ethnic categories were collected into a group “Other”, that was used to help with the numerical estimation of biomarker parameters by estimating the value of prior parameters. The study was restricted to a “healthy” cohort of individuals who had no prior GP record of: stroke or transient ischemic attack (TIA), coronary heart disease (CHD), atrial fibrillation (AF), heart failure (HF), hypertension (HT), peripheral arterial disease (PAD), diabetes mellitus, Asthma, chronic obstructive pulmonary disease (COPD), dementia, Parkinson’s disease, depression, anxiety, serious mental illnesses (bipolar disorder and schizophrenia), cancer excluding non-melanoma skin cancers, chronic kidney disease (CKD), osteoporosis, and rheumatoid arthritis (RA). We also excluded individuals with any prior hospital diagnosed outcomes of: cancer other than non-melanoma skin cancer (ICD-10 coded with C, other than C44), cardiovascular disease (ICD-10 code starting with I), kidney disease (ICD-10 codes N17 or N18), or diabetes (E10-E14). For survival analyses, individuals were censored at any of the hospital diagnosed outcomes (other than the one being studied), or a primary care diagnosis of diabetes. The Supplementary Material SM Table 1, describes the cohort’s formation in terms of numbers of individuals, and Table 1 of the main text describes the resulting cohort, with their number of biomarker measurements and disease outcomes between the start and end of the study period.

**Table 1:** Demographic data of the cohort (ages 45-84 years, with non- or current-smoker status)

| Age Group (years) | 45 to 64 |  | 65 to 84 |  | Disease outcomes |  |  |  |
| --- | --- | --- | --- | --- | --- | --- | --- | --- |
| Sex | Female | Male | Female | Male | Mortality | Ath. dis. | HF | AF |
| Totals | 47240 | 48466 | 22902 | 17373 | 10900 | 9545 | 797 | 2209 |
| Smoking status: never | 36178 | 31840 | 19303 | 13029 | 6776 | 6216 | 574 | 1762 |
| Smoking status: current | 11062 | 16626 | 3599 | 4344 | 4124 | 3329 | 223 | 447 |
| BMI: [18.5,30) | 28367 | 29025 | 13934 | 11236 | 6385 | 5610 | 425 | 1310 |
| BMI: [0,18.5) | 637 | 320 | 400 | 162 | 231 | 82 | 10 | 22 |
| BMI: [30,Inf) | 7348 | 6282 | 2869 | 1625 | 1303 | 1308 | 154 | 332 |
| BMI: Unknown | 10888 | 12839 | 5699 | 4350 | 2981 | 2545 | 208 | 545 |
| Ethnicity: WhiteUK | 40197 | 41315 | 20874 | 15765 | 10372 | 8568 | 709 | 2136 |
| Ethnicity: Asian | 4082 | 3913 | 1212 | 917 | 321 | 717 | 54 | 46 |
| Ethnicity: Black | 2961 | 3238 | 816 | 691 | 207 | 260 | 34 | 27 |
| Socio-economic status: 1 | 8981 | 8503 | 4385 | 3299 | 1656 | 1588 | 115 | 425 |
| Socio-economic status: 2 | 10426 | 9811 | 4994 | 3796 | 2136 | 1904 | 148 | 477 |
| Socio-economic status: 3 | 9761 | 9757 | 4581 | 3592 | 2185 | 1937 | 167 | 495 |
| Socio-economic status: 4 | 9599 | 10539 | 4787 | 3527 | 2334 | 2071 | 185 | 411 |
| Socio-economic status: 5 | 8434 | 9824 | 4135 | 3145 | 2587 | 2040 | 182 | 398 |
| Mortality | 1336 | 2074 | 3674 | 3816 |  |  |  |  |
| Atherosclerotic diseases | 1533 | 3621 | 2101 | 2290 |  |  |  |  |
| Heart failure (HF) | 98 | 120 | 328 | 251 |  |  |  |  |
| Atrial fibrillation (AF) | 399 | 671 | 649 | 490 |  |  |  |  |

### Smoking status, BMI, and missing data

Because of the strong influence of smoking status on health outcomes, it was decided that for this proof-of-principle study, we would exclude individuals for whom smoking status at the start of the study was either unknown or involved past smoking. This ensures that smoking status at the start of the study period is either never smoked, or current smoker. For BMI we have an “unknown” category for individuals whose BMI is unknown.

### Biomarkers

We considered biomarkers that are routinely measured and thought to be associated with cardiovascular health. These included systolic blood pressure (SBP), diastolic blood pressure (DBP), pulse, potassium, sodium, total cholesterol to HDL ratio (TC/HDL ratio), urine albumin to creatinine ratio (UrACR), and C-reactive protein (CRP). To allow the same analysis code to be applied to different biomarkers, the biomarkers of C-reactive protein (CRP) and urine albumin to creatinine ratio (UrACR) were log-transformed to be approximately normally distributed, and all biomarkers were then standardised by subtracting their mean and dividing by their standard deviation before parameters were estimated for each population stratum (as detailed below).

The modeling then proceeds in steps:

1. The (eight) biomarker datasets are split into 192 strata based on age groups (two), ethnic groups (three), sex (two), and smoking status (two).
2. For each stratum, parameters are estimated using a sample of 500 individuals and a sequential Monte Carlo method with a gradient-based maximum likelihood search, applied to a hidden Markov model (HMM) that assumes parameters are the same for everyone in that (comparatively) homogeneous stratum.
3. For each individual, sequential Monte Carlo (SMC) and an expectation-maximisation algorithm was used to estimate their parameters, using a prior parameter estimate from the stratum they belong to and a covariance estimated from all strata. The parameters characterise biomarker trajectories by a mean, a linear slope with age, a within-person variance, and a variance due to short time-scale variations or measurement error.
4. The estimated parameters were split into tertiles. This allowed an “unknown” category to model individuals whose biomarker data were too few (less than three) or non-existent, and ensured the analysis would be more robust to noise or outliers that might arise from the SMC estimates. Associations between individual outcomes and within-person biomarker variances were estimated with a proportional hazards model, that was stratified by age-group and adjusted for ethnicity, smoking status, BMI category, and socio-economic status.

Between steps 3 and 4, histograms of individuals’ estimates were inspected and as a “quality control” precaution against potentially unreliable estimations, five strata with an estimate more than three standard deviations from the mean were removed. Details of the HMM are in the Supplementary Material. The results of the proportional hazard fits are shown in forest plots, with additional plots in the Supplementary Material that find similar results, but simultaneously adjust for both mean biomarker values and standard deviations. Note that in the forest plots each biomarker association is estimated independently of the other biomarkers.

## Results

We consider hospital-diagnosed outcomes of: mortality (figure 1), atherosclerotic diseases (ICD-10 codes I20-25, I63-70, figure 2), heart failure (ICD-10 code I50, figure 2, and atrial fibrillation (ICD-10 code I48, figure in Supplementary Material). Associations are considered with tertiles of the within-person variances of the eight biomarkers: total cholesterol to HDL ratio (TC/HDL ratio), systolic blood pressure (SBP), diastolic blood pressure (DBP), Pulse, potassium, sodium, urine albumin to creatinine ratio (UrACR), and C-reactive protein (CRP). Associations are estimated with a proportional hazards model as described in the Methods, with stratification by age group and adjustment for: smoking status, BMI category, sex, ethnicity, and socio-economic status. The baseline was taken as “unknown”, although to allow visual comparison of the estimated associations, the fits are offset so that the lowest tertile is centered at 1. Each figure plots three columns, using biomarker data that is considered either: up to the end of study period or an outcome (righthand column), 2-years prior to the end of study period or an outcome (left-hand column), or 1-year prior to the end of study period or an outcome (middle column). There are (reassuringly) small differences between the three columns of results, with the greatest differences between estimates using biomarker data up to 0-years prior to the study’s end, and data up to 1- or 2-years before the end of the study. The associations are strongest with (all cause) mortality, with the weakest associations in that case being with UrACR and CRP. As can be seen in table 2, there are far fewer measurements of UrACR and CRP than the other biomarkers, explaining the much larger confidence intervals and uncertainty over their strength of association. The other biomarker associations are clearer, typically with relative risks of 1.1 to 1.2, and with a dose-response relationship leading to greater within-person variances being associated with systematically greater relative risks (or vice versa). A clear association and a dose-response relationship, both strengthen the evidence for a causal interpretation of the associations [53].

**Table 2:** Number of individuals with biomarker measurements, and disease outcomes.

| Biomarker | Mortality | Disease outcomes |  |  |
| --- | --- | --- | --- | --- |
|  |  | Atherosclerotic diseases | Heart failure | Atrial fibrillation |
| TC/HDL ratio | 3060 | 2729 | 251 | 697 |
| SBP | 6036 | 4500 | 476 | 1150 |
| DBP | 5959 | 4450 | 472 | 1138 |
| Pulse | 1491 | 912 | 135 | 280 |
| Potassium | 4288 | 2989 | 369 | 791 |
| Sodium | 4326 | 3011 | 370 | 802 |
| UrACR | 288 | 122 | 25 | 34 |
| CRP | 515 | 366 | 55 | 108 |

**Figure 1:**
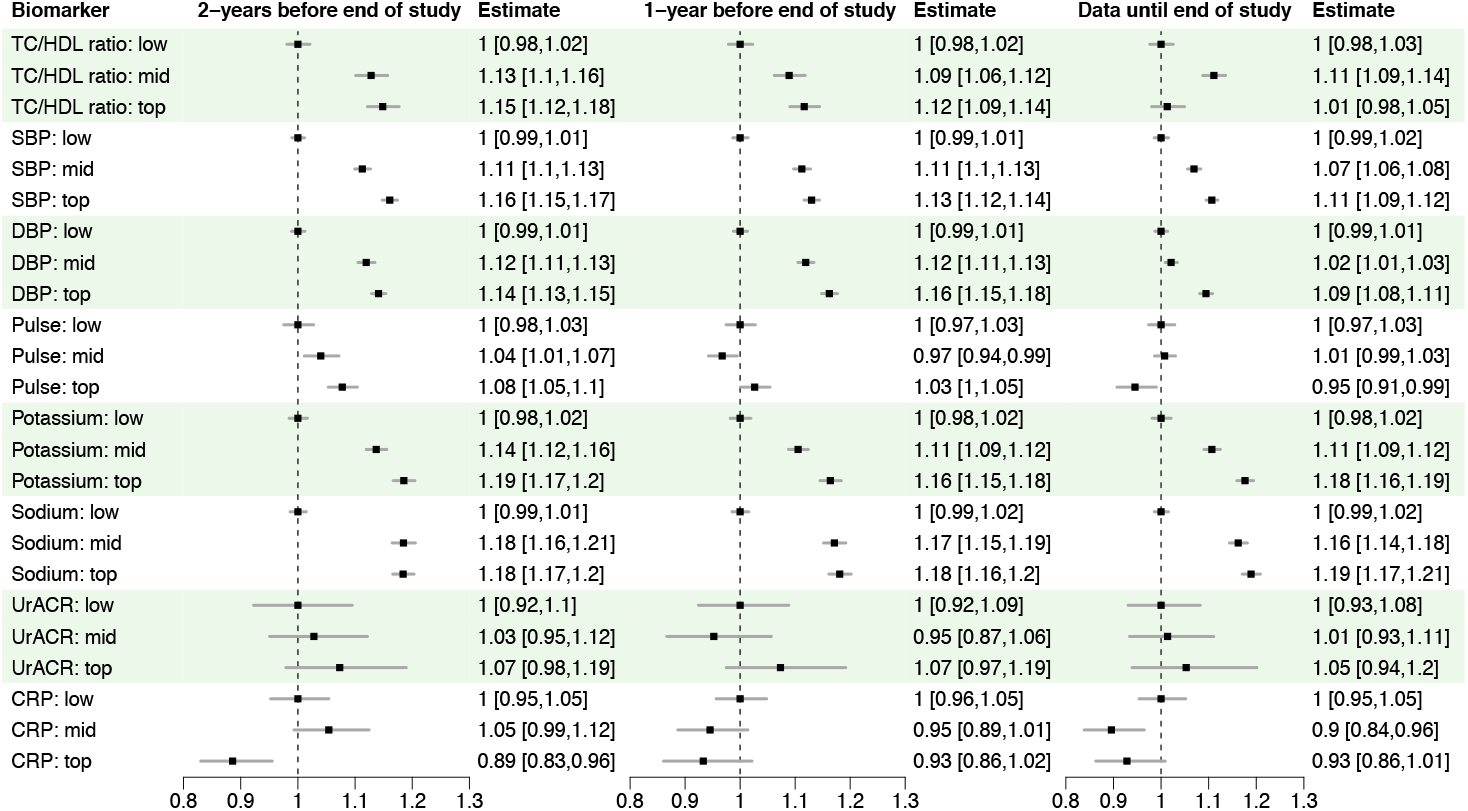
Associations between mortality and within-person biomarker standard deviation, stratified by age-group, adjusted for sex, ethnicity, BMI tertile, smoking status, and socio-economic status. For easier visual comparison, plots offset the lowest tertile to 1. Fits use biomarker data until either of a CVD diagnosis, censoring, or end of follow-up period (right), data up to 2-years before this (left), and data up to 1-year before this (middle). Studies with adjustment for mean biomarker value are in the Supplementary Material, with minor differences.

**Figure 2:**
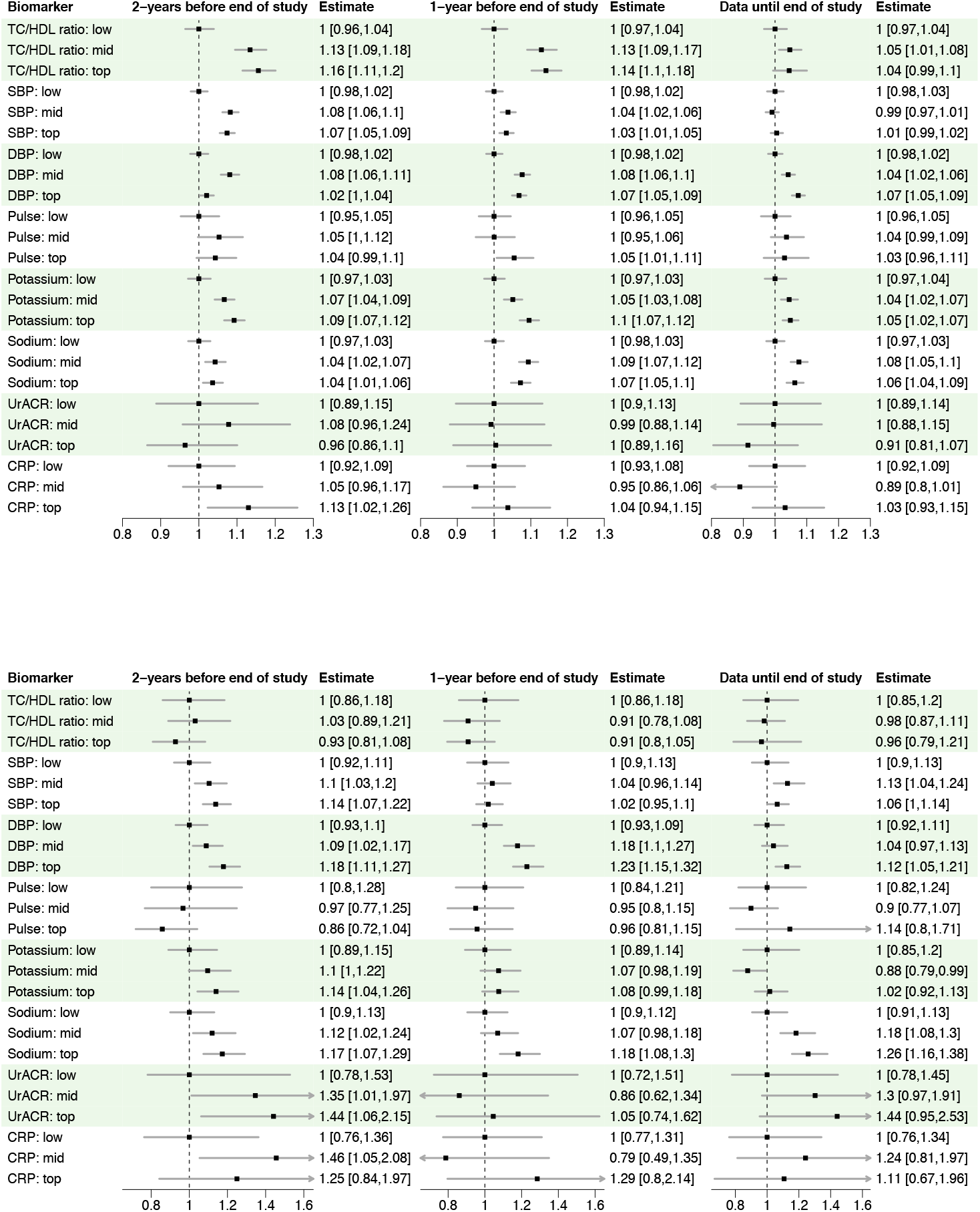
Top: Associations between atherosclerotic heart diseases (ICD-10 codes: I20-25, I63-70), and within-person biomarker standard deviation. Bottom: Associations between heart failure (ICD-10 code I50), and within-person biomarker standard deviation. Both studies are stratified by age-group, adjusted for sex, ethnicity, BMI tertile, smoking status, and socio-economic status. For easier visual comparison, plots offset the lowest tertile to 1. Fits use biomarker data until either of a CVD diagnosis, censoring, or end of followup period (right), data up to 2-years before this (left), and data up to 1-year before this (middle). Studies with adjustment for mean biomarker value are in the Supplementary Material, with minor differences.

Equivalent studies with simultaneous adjustment for mean biomarker value are in the Supplementary Material. These more complex plots need more space to display and effort to interpret, but they show similar results, with minor strengthening or weakening of associations that depended on the tertile of the mean biomarker’s estimate. Although in the Supplementary Material, these results are critical, because they indicated that associations with within-person biomarker variability were independent of associations with biomarker level.

Sensitivity analyses compared the estimated biomarker means and within-person variances, with the number of measurements that an individual has (see Supplementary Material). Any associations with the number of measurements were small when compared with the standard deviation of the estimates, that varied far more between individuals with the same number of measurements, than on average between individuals with different numbers of measurements. This observation, and the comparative insensitivity to the time prior to the end of study over which biomarkers are measured, support the suggestion that it may be possible to avoid or neglect some of the inherent biases that may arise from associations between an individual experiencing symptoms and biomarker measurements being made. Supplemental plots also supported the hypothesis that a more extreme biomarker observation, would be associated with additional measurements to confirm whether it was incorrect, or temporary (Supplementary Material).

## Discussion

The results show clear associations between individuals’ within-person biomarker variances and disease risks. When we simultaneously adjusted for the variance and mean biomarker values, these associations remained and sometimes became stronger (see Supplementary Material). This is despite potential inaccuracies of biomarker estimates, and the subsequent choice to group these into tertiles. Two serious concerns we had were: whether the strength of associations were sensitive to the proximity of disease onset, and whether biomarker estimates were sensitive to the number of data measurements that an individual had. Figures 1–2 indicated that associations are only weakly influenced by the proximity to disease onset, and suggest that a gap of one or two years is probably sufficient to ensure that estimates from association studies are unaffected by disease onset. Regarding our second concern, results in the Supplementary Material show that correlations between biomarker estimates and the number of data measurements are weak compared with the differences between individual estimates. Together, these sensitivity studies suggest that neither of these concerns will prevent the use of this approach for improving disease risk identification.

### What is the mechanism?

The clear associations and dose-response relationship - with stronger (weaker) associations for the max (min), or vice versa, add support to a potential causal relationship between the biomarker variances and disease risk. However the associations between biomarker variances and disease risk are not yet clearly explained in terms of causal mechanisms. The “E-value” [53] assesses the strength of confounding needed to explain the observed risk-factor association. An estimated relative risk of ~1.2 can be explained by a confounding factor whose risk ratio is at least ~1.7 or greater [53]. So an explanation based on unmeasured confounding is possible, but requires a comparatively strong unidentified confounder that is associated with both disease risk and biomarker variance.

Biomarker variances have been linked with potentially disease-promoting biological mechanisms. Blood pressure variation has been associated with changes in the p-wave of the heart [19]. It has also been linked with inflammation and endothelial growth factors [20–22], that both offer a potential underlying cause and mediating factor linking blood pressure variability to disease risks. Possible mechanisms linking blood pressure variation and disease are discussed further in [23]. Potential causal mechanisms for the role of LDL variability in disease risk have also been postulated [25], with mechanisms that include increased plaque instability [54] and endothelial dysfunction [55]. In general however the role of biomarker variation is poorly understood. It remains to be determined whether biomarker variation represents a symptom of an underlying condition, a direct cause of disease risk, or a combination of both.

### Impediments to practical implementation

The research here demonstrates the potential importance of biomarker variability for assessing disease risk, and provides a proof-of-principle methodology for its estimation using routinely measured biomarkers. Because the biomarker parameters are estimated with routinely collected GP data, we have also demonstrated the potential for improving the early identification of disease through improved analyses of existing data.

There are at least two impediments to incorporating these results into disease-risk models. One is to generalise the method so that it can be used with individuals that are not in one of the population strata studied here. This should be comparatively easy to do. For example, we could match the individual to 1000 similar people, and use them to provide a prior estimate for biomarker parameters, and then continue as before with individual parameters estimated using the prior and the individual’s data.

A greater challenge is to account for the strongly informative presence, missingness, and timing of biomarker measurements. A striking result from the study here was the strong association between the existence of a measurement and subsequent disease risk. These associations are complex, and it is likely that future work will need to simultaneously model the timing of measurements, their value, and disease risk. A quick fix is to turn this problem into an opportunity, and cluster individuals based on the presence and value of measurements, and form population subsets with similar risk. Difficulties with this approach are that it requires work to interpret the meaning of the clusters, and it would provide a crude approximation to a more complete statistical model. An alternative is to model the decision to make a measurement, and to do so sufficiently well that it can account for the information inherent in knowing that a measurement has been made.

### Next steps

There is scope to develop improved estimates for biomarkers such as blood pressure and blood pressure variability. For example we might imagine a system where a doctor inputs a patient’s details (age, sex, ethnicity, BMI, smoking status etc), to a remote computer with access to a large database of people. A remote calculation would then extract a sample of 1000 similar individuals from which to estimate a set of prior biomarker parameters, that would then be combined with the individual’s biomarker data to estimate their biomarker’s mean, slope, and within-person variation.

Here biomarker trajectories and disease risk were modeled separately. The advantage of using a secondary modeling procedure for disease risk is that it allowed conventional methods to be used, and provided results in a form that are familiar to most epidemiologists. However, the uncertainty in the estimated parameters was not accounted for in estimating effect sizes, which could lead to regression dilution bias. Furthermore, it is unclear how to interpret associations with multiple strongly correlated measurements if these are included simultaneously into a disease-risk model.

Ideally, many of an individual’s biomarker measurements would be used to improve the performance of disease-risk models. This requires modeling all of the potentially important information, including the fact that a measurement has been taken. It is highly informative that an individual has visited their doctor, and that the doctor has decided to make a series of measurements. These may be needed to systematically rule out potential underlying causes, and are likely to produce a highly correlated selection of measurements that are determined by current medical practice. This makes combining measurements into a multivariate model challenging, and seems likely to need a statistical model that combines the decision to make a measurement, the values that are obtained, and possibly also includes records of disease diagnoses or underlying conditions.

In addition to these key challenges, it seems likely that an improved mathematical model for the unmeasured latent trajectories can be found. Ideally the latent model would reflect biology in a more informative way than the “random walk” model that is used here to reflect our ignorance of the underlying processes. The random walk model has successfully described a wide variety of systems, but there is scope for a less generic model that either performs better, or is able to provide insights into underlying biological changes.

## Conclusions

We have estimated associations between the within-person variation of eight biomarkers and risk of mortality or cardiovascular disease, in a cohort of 45-84 year old individuals of Black, White UK, and Asian (non-Chinese) ancestry, who at the study’s start had no previous report of cardiovascular disease or cancer (other than non-melanoma skin cancers, ICD-10 code C44). Clear associations were found with TC/HDL ratio, SBP, DBP, Pulse, Potassium, and Sodium, and weaker evidence of associations can be seen with UrACR and CRP, for which fewer data were available. These associations remained when simultaneously adjusted for mean biomarker values. These results must now be translated into robust scientific code that can be implemented in real-time in a general practitioner setting with a wider range of biomarkers, and augmented with a rigorous scientific methodology to quantify the link between biomarker trajectories and disease risk. The results here vindicate this approach, demonstrating the potential importance of within-person biomarker variance for assessing disease risk, and providing a proof-of-principle demonstration that routinely measured biomarkers can be used for their estimation.

## Author contributions

Conceptualization and project administration: The trajectories component of the COMPUTE project was conceived and administered by RP and DS. The methodology was developed by AW and DS, with analysis, coding, visualisation, and initial drafts by AW. Data acquisition and curation was by CWD and the Primary Care COMPUTE team. All authors read and commented on the final draft.

## Data availability

Data are available on application from the Clinical Practice Research Datalink (CPRD).

https://www.cprd.com/

## Code availability

The R code [56] used to produce figures will become available after publication at: https://osf.io/

R packages used in this study include survival [57], grr [58], data.table [59], and xtable [60].

## Footnotes

1 Citations for sodium and potassium exclude patients on dialysis.

